# Early Awake Prone and Lateral Position in Non-intubated Severe and Critical Patients with COVID-19 in Wuhan: A Respective Cohort Study

**DOI:** 10.1101/2020.05.09.20091454

**Authors:** Wei Dong, Yiping Gong, Juan Feng, Lang Bai, Haomiao Qing, Peng Zhou, Yu Du, Junchen Zhu, Shanling Xu

## Abstract

**Background:** Previous studies suggest applying prone position (PP) and lateral position (LP) in patients with severe acute respiratory distress syndrome (ARDS) for their efficacy in improving oxygenation and lung recruitment.This paper aims to share clinical experiences and outcome of using PP and LP in combination with oxygen therapy (OT) and Non-invasive ventilation (NIV) in severe and critical patients with COVID-19.

**Methods:** Clinical data of 48 severe and critical patients have been retrieved from medical records and reviewed. The primary outcome is the survival rate. Secondary outcome is the rate of patients requiring intubation.

**Results:** In total, 25 patients were finally included in the study. The mean respiratory rate of all 25 patients decreased from 28.4 breaths/min to 21.3 breaths/min. CT results showed increase in lung recruitment. All patients tolerated PP and LP well. No deterioration or severe adverse events associated with PP and LP occurred. All patients recovered and survived without intubation. Follow-up to date showed that all patients have been discharged except one with mild symptoms and positive RNA test.

**Conclusion:** Clinical outcomes of early application of PP and LP in combination with OT and NIV in severe and critical patients with COVID-19 indicated well tolerance of the therapy and resulted in improving patients’ oxygenation in a safe and effective manner. Therefore, this strategy can be explored as an early intervention in managing patients in early stage of disease development under the context of pandemic and limited medical resources.

## Introduction

The outbreak of a novel coronavirus disease (COVID-19) started in Wuhan, China in December, 2019 and has since rapidly impacted many countries as a pandemic. According to WHO’s COVID-19 situation report on May 4, 2020, there are nearly 3.44 million confirmed cases worldwide, with 239,604 deaths.^1^ According to the findings in China, about 14% of COVID-19 patients were severe and 5% were critical and the overall case-fatality rate among critical cases reached 49.0%.^2^ These findings have greatly stressed the health care system globally. So far there is no specific antiviral drugs for the treatments of COVID-19. Whilesome studies^3-6^ have reported on the clinical characteristics and outcomes of critically ill patients, the challenges of managing these patients remain particularly in the context of limited medical resources and the global surge of patients. Evaluating the effects of supportive therapies becomes one of the priorities of research.^7^

Current Chinese clinical management guidelines^8^ for COVID-19 suggest treatment including rest in bed, oxygen therapy, antiviral medication, and mechanical ventilation. For critical patients with severe ARDS, many guidelines^8-11^ recommend prone ventilation for 12-16 hours a day, especially for adult patients. This is in alignment with previous Randomized Controlled Trials (RCTs) and studies^12,13^ that showed the impact of prone position in improving oxygen and lung recruitment in mechanically ventilated patients with severe ARDS.

Based on previous clinical experiences in managing patients with severe acute respiratory infection, we used early prone position (PP) and lateral position (LP) in combination with oxygen therapy (OT) and non-invasive ventilation (NIV), as supportive therapies, to treat severe and critical patients with COVID-19. We found these therapies to be effective interventions in early management of patients with critical and/or severe symptoms of COVID-19. The aim of this study is to share the clinical characteristics and outcomes of severe and critical patients using these interventions. The primary outcome is the survival rate of the patients. Secondary outcome is the rate of patients requiring invasive mechanical ventilation.

The study was approved by the Institutional Review Board (IRB) for clinical research at Renmin Hospital of Wuhan University (WDRY2020-K125). Written informed consent was waived as this is a designated hospital for treatment of COVID-19 patients.

## Methods

### Study Design and Participants

This observational, retrospective study was completed at Renmin Hospital of Wuhan University, one of the designated hospitalsin Wuhan to treatpatients with COVID-19. In late January, 2020, with the rapid increase of patients in Wuhan, medical teams from Sichuan Province were mobilized and sent to Wuhan to support local hospitals. On February 5, 2020, our team took over Ward No. 6 (a temporary ICU). Between February 5 and February 29, 2020, there were a total of 48 severe and critical patients with laboratory-confirmed diagnosis of COVID-19. Severe and critical patients were defined according to the Chinese guidelines^8^. Patients with respiratory distress (RR≥30 breaths/min), or SpO2≤93% at rest, or PaO2/FiO2≤300mmhg were defined severe; while patients with respiratory failure requiring mechanical ventilation, or shock, or other organ failure requiring intensive care were critical. All 48 patients received standard treatment including antiviral medication, antibiotics when necessary, anticoagulation and nutritional support. Oxygen therapy using different devices including nasal prongs (NP), mask, high flow nasal cannula (HFNC) was provided to all patients based on their status of oxygenation. Non-invasive ventilation was provided based on patients’ needs. Humidification was also added for all patients.

In addition, for patients with PaO2/FiO2 <200, or whose CT results indicated acute exudation of both lungs, a daily PP session of more than 10 hours (PP session>10 hours/day) was ordered. For other patients, a daily PP session of more than 4 hours (PP session>4 hours/day) was ordered. Nurses provided instructions to patients as how to perform proper PP. Changes in vital signs, adverse events, after PP were observed and documented by the nurses. No sedation was provided. Actual hours of PP sessions were self-reported by the patients dailyand further checked and documented by nurses. For patients who did not tolerate PP sessions as ordered, a combination with PP and LP, or LP only was suggested instead. In addition, for critical patients with HFNC or NIV support, audits by nurses or peer patients were encouraged to ensure compliance.

### Data Collection

All the medical and nursing records, laboratory findings, and radiological examinations for those 48 patients have been reviewed including information during the clinical course, from onset, admission to the hospital, to admission to unit. Demographic, treatment, and outcome data have been collected and analyzed.

### Analysis

The aim of this observational study is to share the clinical outcomes of severe and critical patients treated with PP and LP in combination with OT and NIV as supportive therapies, no hypothesis as in the RCTs has been designed or implemented to include certain sample sizes. We have included all available cases on the unit.

Patients were divided into severe and critical groups based on severity and comparison was made between the two groups. Statistical analyses were conducted using SAS statistical software (version 9.3). The continuous variables are presented as mean ± standard deviation for normally distributed data or median ± interquartile range for non-normal data, and the categorical variables are reported as number (percentage). The change of respiratory rate (RR), CT score before and after the PP and LP, were carried out using paired t-tests.

CT was evaluated by a semi-quantitative score system that assessed each segment of the lobes for the degree of segmental involvement and abnormalities. The scoring is 1) No involvement score 0; involvement<1/3 to a lobe score 1, involvement≥ 1/3 and ≤2/3 to a lobe score 2; involvement>2/3 to a lobe score 3; 2) abnormalities: no abnormality score 0; major abnormality as ground glass opacity (GGO) score 1; major abnormality as consolidation as major abnormality score 2. For each segment, the score is from 1 to 5. In total, the maximum total score for two lungs is 100. Two senior radiologists with more than 5 years of working experience evaluated the images independently according to the scoring system and the mean score of the two was accepted as final. For any difference of higher than 5 scores between two evaluations, the images were reviewed by a third radiologist and a mutually agreed score will be accepted thereafter.

## Results

Among the 48 patients, 2 were excluded due to an early transfer for non-medical reasons within 5 days. Another 8 critical patients were excluded due to failure in PP nor LP without intubation. Among them, 1 ended up using invasive mechanical ventilation and prone position, and survived. The other 7 received NIV and died due to disease progression. In total, 38 patients used PP and/or LP in combination with oxygen therapy and NIV. However,13 of them quickly changed from severe to mild within 2-5 observation days and got discharged, therefore were excluded in this study. Finally 25 patients were enrolled, including 12 severe and 13 critical. (Figure 1). The clinical charts of adult patients with laboratory-confirmed COVID-19 admitted in the ICU were reviewed. The mean age was 49.0±14.1 years in the severe group, while 59.5±16.7 years in the critical group (Table 1). 9 (36%) have comorbidities. More than 50% of patients have hypercapnia. Less than 50% of patients had a decrease in CD4+ and CD8+ and abnormal D-dimer test results. For the Acute Physiology and Chronic Health Evaluation II (APACHE II) score that is greater than 15, there were 3 in severe group and 11 in critical group. While for the median Sequential Organ Failure Assessment (SOFA) score that is greater than 2, there were 6 in severe group and 11 in critical group. All patients were provided antiviral treatment, while 14 (56%) were treated with antibiotics and 12 (48%) were treated with glucocorticoid. All patients received oxygen therapy, with 10 (40%) used NP, 6 (24%) simple face mask, and 6 (24%) HFNC. 3 (12%) patients required NIV support. Among 25 patients, 21 tolerated a daily PP session for 1-14 hours (Table 2, Table 3). Out of the 21 that tolerated a daily PP session, 6 tolerated a daily PP session less than 2 hours, and added a daily LP for 4 hours and 4 only tolerated a daily LP session for 4 hours (all of older ages, 56, 78, 79 and 82 years respectively). During the study period, the mean daily PP session was 6.4±3.7 hours in critical group, and 3.6±1.6 hours in severe group. 9 had adverse events after PP and/or LP session, including 4 with dyspnea and sternal pain, 1 scrotal pain, 1 with lumbago that was relieved after adjusting the position, and 1 with itchiness at chest wall skin.

**Figure 1:**
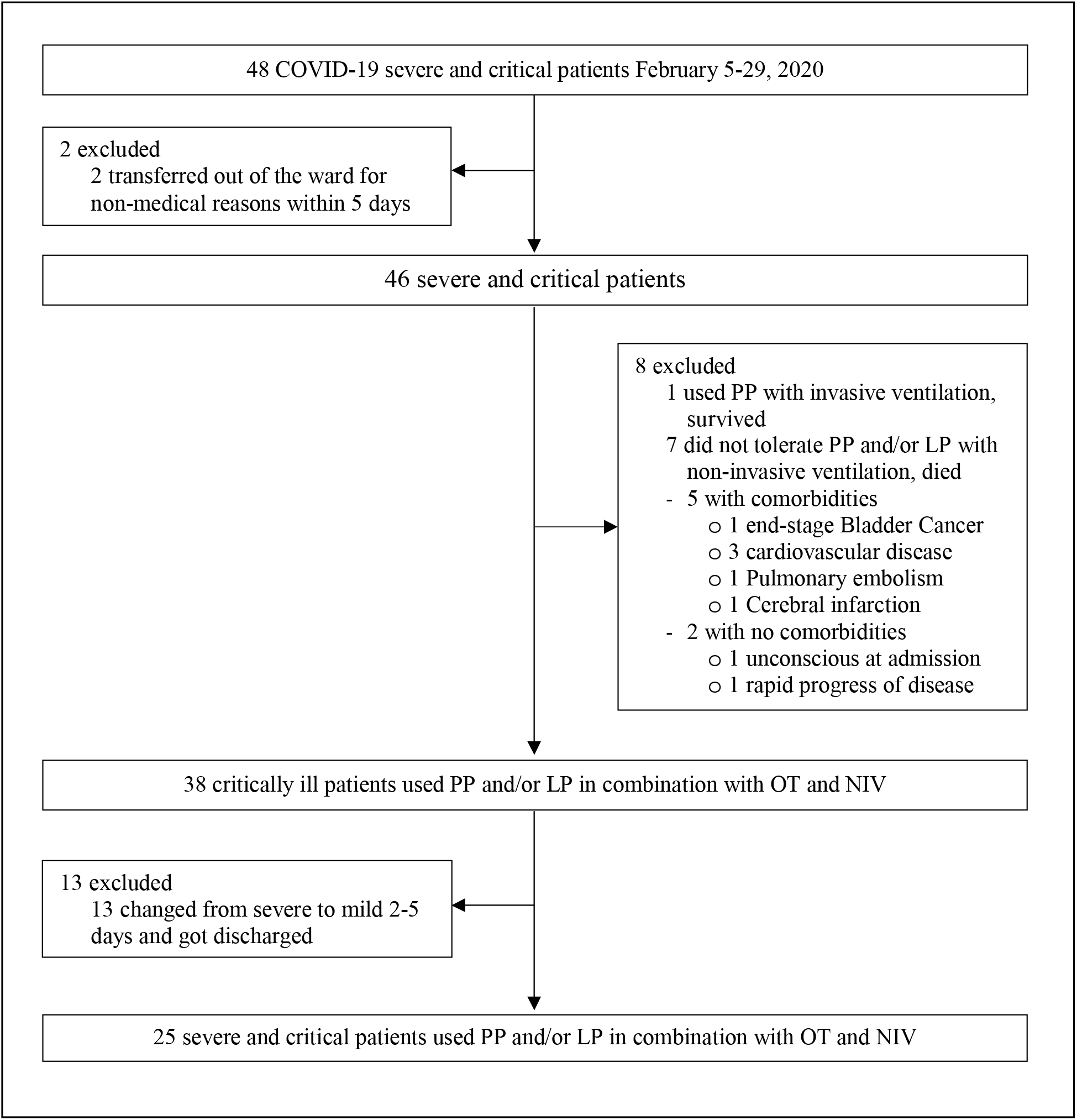
Study flow Diagram

**Table 1.** Clinical characteristics of the study patients

| Characteristic | All Patients<br>(N=25) | Disease Severity |  |
| --- | --- | --- | --- |
|  |  | Severe<br>(N=12) | Critical<br>(N=13) |
| Age - n(%) |  |  |  |
| Mean | 54.4(16.1) | 49.0(14.1) | 59.5(16.7) |
| Distribution |  |  |  |
| 20-40 | 6 (24.0) | 3 (25.0) | 3 (23.1) |
| 41-64 | 14 (56.0) | 8 (66.7) | 6 (46.2) |
| ≥65 yr | 5 (20.0) | 1 (8.3) | 4 (30.8) |
| Sex - n(%) |  |  |  |
| Male | 16 (64.0) | 5 (41.7) | 11 (84.6) |
| Female | 9 (36.0) | 7 (58.3) | 2 (15.4) |
| Smoking history - n(%) | 7(28.0) | 4 (33.3) | 3 (23.1) |
| Family Cluster History - n(%) | 18 (72.0) | 9 (75) | 9 (69.2) |
| Symptoms - n(%) |  |  |  |
| Fever | 25(100) | 12(100) | 13(100) |
| Median highest temperature - °C | 38.8 (1.3) | 38.6 (1.1) | 39.0 (1.5) |
| 37.5-38.0 °C | 8 (32.0) | 4 (33.3) | 4 (30.8) |
| 38.1-39.0 °C | 8 (32.0) | 5 (41.7) | 3 (23.1) |
| >39.0 °C | 9 (36.0) | 3 (25.0) | 6 (46.1) |
| Cough | 21 (84.0) | 11 (91.7) | 10 (76.9) |
| Sputum production | 11 (44.0) | 5 (41.7) | 6 (46.2) |
| Dyspnea | 25 (100.0) | 12 (100) | 13 (100) |
| Fatigue | 15 (60.0) | 7 (58.3) | 8 (61.5) |
| Diarrhea | 1 (4.0) | 0 (0) | 1 (7.7) |
| Poor Appetite | 7 (28.0) | 4 (33.3) | 3 (23.1) |
| Myalgia | 4 (16.0) | 2 (16.7) | 2 (15.4) |
| Headache | 5 (20.0) | 4 (33.3) | 1 (7.7) |
| Any Comorbidity - n(%) |  |  |  |
| Any | 9 (36) | 4 (33.3) | 5 (38.5) |
| Hypertension (HTN) | 6 (24) | 3 (25) | 3 (23.1) |
| Diabetes | 1 (4.0) | 0 (0) | 1 (7.7) |
| Gout | 1 (4.0) | 0 (0) | 1 (7.7) |
| Chronic Hepatitis B (CHB) | 1 (4.0) | 0 (0) | 1 (7.7) |
| Gastrointestinal stromal tumor (GIST) | 1 (4.0) | 0 (0) | 1 (7.7) |
| Arrhythmia | 2 (8.0) | 1 (8.3) | 1 (7.7) |
| Meniere | 1 (4.0) | 1 (8.3) | 0 (0) |

**Table 2.** Laboratory Data, Baseline characteristics at Ward (ICU) and Treatment

|  | All Patients<br>(N=25) | Disease Severity |  |
| --- | --- | --- | --- |
|  |  | Severe<br>(n=12) | Critical<br>(n=13) |
| **Arterial blood gas analysis\* - n/total n** |  |  |  |
| pH <7.35 and >7.45 | 14/20 | 4/8 | 10/12 |
| PaCO2>45mmHg | 13/20 | 4/8 | 9/12 |
| Lactate > 2mmol/L | 10/20 | 3/8 | 7/12 |
| PaO2/FiO2< 300mmHg | 17/20 | 6/8 | 11/12 |
| **Blood routine- no.(%)** |  |  |  |
| White-cell count>10,000/mm<sup>3</sup> | 3(12) | 0(0) | 3(23) |
| Lymphocyte count<1,500/mm<sup>3</sup> | 17(68) | 5(42) | 12(92) |
| Platelet count <150,000/mm<sup>3</sup> | 4(16) | 1(8) | 3(23) |
| **Blood biochemical analysis- no.(%)** |  |  |  |
| Alanine aminotransferase >40 U/L | 11(44) | 4(33) | 7(54) |
| Aspartate aminotransferase >40 U/L | 8(32) | 4(33) | 4(31) |
| Serum total bilirubin >17.1umol/L | 5(20) | 3(25) | 2(15) |
| Serum creatinine >110umol/L | 1(4) | 0(0) | 1(8) |
| Serum lactate dehydrogenase >250U/L | 8(32) | 5(42) | 3(23) |
| **Others<sup>#</sup>- n/total n** |  |  |  |
| C-reactive protein≥10mg/L- no.(%) | 17(68) | 6(50) | 11(85) |
| Procalcitonin≥0.5ng/ml | 2/22 | 0/10 | 2/12 |
| CD4+ T cells count<400/uL<sup>\$</sup> | 11/24 | 2/11 | 9/13 |
| CD8+ T cells count <220/uL<sup>\$</sup> | 13/24 | 2/11 | 11/13 |
| D-Dimer>0.5mg/L<sup>#</sup> | 16/22 | 6/11 | 10/11 |
| Prothrombin activity<75%<sup>#</sup> | 6/22 | 1/11 | 5/11 |
| Apache II scoreon before PP≥15<sup>&</sup> | 14/20 | 3/8 | 11/12 |
| SOFA scoreon before PP≥2<sup>&</sup> | 17/20 | 6/8 | 11/12 |
| **Treatment** |  |  |  |
| Antiviral- no.(%) | 19(100) | 12(100) | 13(100) |
| Antibiotics- no.(%) | 14(56) | 4(33) | 10(77) |
| Corticosteroids- no.(%) | 12(48) | 2(17) | 10(77) |
| Intravenous immunoglobulin- no.(%) | 19(76) | 7(58) | 12(92) |
| Anticoagulation- no.(%) | 7(28) | 3(25) | 4(31) |
| Oxygen therapy - no.(%) |  |  |  |
| Nasal prongs | 10(40) | 10(83.3) | 0(0) |
| Face mask | 6(24) | 1(8.3) | 5(38.5) |
| High-flow nasal cannula (HFNC) | 6(24) | 1(8.3) | 5(38.5) |
| Mechanical ventilation - no.(%) |  |  |  |
| Noninvasive | 3(12.0) | 0(0.0) | 3(23.1) |
| Positioning- no.(%) |  |  |  |
| PP | 15(60) | 7(58.3) | 8(61.5) |
| LP | 4(16) | 1(8.3) | 3(23.0) |
| PP+ LP | 6(24) | 4(33.3) | 2(15.3) |
| Mean Daily PP session hours<sup>\$</sup> | 4.9(3.1) | 3.6(1.6) | 6.4(3.7) |
| Mean Baseline RR<sup>\$\$</sup> | 28.4(3.5) | 26.6(2.1) | 30.0(3.8) |
| Mean Post RR<sup>\$\$</sup> | 21.3(1.3) | 20.5(1.2) | 22.1(1.0) |
| Mean Baseline CT-score<sup>\$\$</sup> | 41.8(19.2) | 26.7(11.6) | 55.8(13.2) |
| Mean Post CT-score<sup>\$\$</sup> | 29.3(16.8) | 16.4(7.7) | 41.2(13.7) |

\*Data were available for 20 patients
#Data were available for 22 patients

**Table 3.** Treatment and outcomes of the studies patients

| Case No. | Sex | Age | Comorbidities | Severity | Apache II /SOFA | Treatment | Daily PP and/or LP Session (hrs) | Baseline PaO<sub>2</sub>/FiO<sub>2</sub> | Post PaO<sub>2</sub>/FiO<sub>2</sub> (days*) | Baseline RR | Post RR (days*) | Baseline CTscore | Post CT score (days*) | Adverse events Post PP/LP | Outcome |
| --- | --- | --- | --- | --- | --- | --- | --- | --- | --- | --- | --- | --- | --- | --- | --- |
| 1 | Male | 43 | HTN,CHB | Critical | 15/2 | HFNC+PP | 8 | 222 | 295(5) | 30 | 22(6) | 65 | 47(7) | Scrotal pain | Survive |
| 2 | Male | 79 |  | Critical | 23/3 | HFNC+LP | 4 | 198 | 348(17) | 29 | 23(1) | 56 | 52(10) | Dyspnea, Sternal pain | Survive |
| 3 | Male | 60 | CC | Critical | 18/1 | HFNC+PP | 6 | 330 | 443(4) | 28 | 20(6) | 25 | 17(6) | Sternal pain | Survive |
| 4 | Female | 62 |  | Critical | 15/4 | HFNC+PP+ LP | 2+4 | 166 | 209(6) | 30 | 23(2) | 66 | 47(7) |  | Survive |
| 5 | Male | 59 | HTN | Critical | 25/3 | HFNC+PP | 10 | 123 | 363(7) | 27 | 22(5) | 46 | 36(8) | Skin itchiness | Survive |
| 6 | Female | 79 |  | Critical | 18/3 | NIV+PP+ LP | 2+4 | 128 | 184(4) | 31 | 21(7) | 53 | 22(13) | Chest tightness | Survive |
| 7 | Male | 56 | Arrhythmia | Critical | 24/5 | NIV+PP | 8 | 162 | 392(6) | 33 | 23(6) | 55 | 43(6) |  | Survive |
| 8 | Male | 36 |  | Critical | 23/3 | NIV+PP | 14 | 184 | 418(3) | 40 | 23(7) | 80 | 45(12) |  | Survive |
| 9 | Male | 39 |  | Critical | Missing | Mask+PP | 4 | Missing | 436 | 33 | 22(3) | 56 | 49(10) |  | Survive |
| 10 | Male | 38 |  | Critical | 12/3 | Mask+PP | 6 | 178 | 373(7) | 26 | 22(7) | 61 | 60(8) |  | Survive |
| 11 | Male | 78 | HTN,DM,Gout | Critical | 30/5 | Mask+LP | 4 | 162 | 390(9) | 27 | 22(5) | 57 | 31(13) | Dyspnea, Sternal pain | Survive |
| 12 | Male | 82 |  | Critical | 18/4 | Mask+LP | 4 | 181 | Missing | 26 | 21(4) | 42 | 27(12) | Dyspnea, Sternal pain | Survive |
| 13 | Male | 62 |  | Critical | 18/3 | Mask+PP | 4 | 158 | 388(15) | 30 | 23(4) | 63 | 60(11) | Lumbago | Survive |
| 14 | Female | 65 | HTN,Meniere | Severe | 18/3 | HFNC+PP+ LP | 2+4 | 190 | 229(7) | 31 | 22(6) | 50 | 28(11) |  | Survive |
| 15 | Male | 63 |  | Severe | Missing | Mask+PP | 4 | Missing | Missing | 26 | 18(3) | 28 | 19(13) |  | Survive |
| 16 | Male | 42 |  | Severe | 8/2 | NP+PP+ LP | 2+4 | 224 | 306(7) | 28 | 22(4) | 19 | 13(6) |  | Survive |
| 17 | Male | 32 |  | Severe | Missing | NP+PP+ LP | 2+4 | Missing | Missing | 26 | 22(2) | 46 | 22(10) |  | Survive |
| 18 | Male | 29 |  | Severe | Missing | NP+PP+ LP | 1+4 | Missing | Missing | 25 | 20(5) | 23 | 6(13) |  | Survive |
| 19 | Female | 61 | HTN | Severe | Missing | NP+PP | 6 | Missing | Missing | 24 | 20(7) | 32 | 25(12) |  | Survive |
| 20 | Female | 56 | HTN | Severe | 16/2 | NP+PP | 4 | 290 | Missing | 25 | 19(2) | 29 | 25(8) |  | Survive |
| 21 | Female | 41 |  | Severe | 7/2 | NP+PP | 4 | 215 | 312(7) | 26 | 21(3) | 11 | 9(7) |  | Survive |
| 22 | Female | 27 |  | Severe | 12/0 | NP+PP | 4 | 415 | Missing | 27 | 21(3) | 15 | 6(7) |  | Survive |
| 23 | Female | 53 |  | Severe | 9/2 | NP+PP | 4 | 232 | 288(9) | 26 | 20(5) | 25 | 15(7) |  | Survive |
| 24 | Female | 63 |  | Severe | 15/2 | NP+PP | 6 | 273 | 366(2) | 25 | 20(1) | 18 | 10(7) |  | Survive |
| 25 | Female | 56 | Arrhythmia | Severe | 13/0 | NP+ LP | 4 | 420 | Missing | 30 | 21(7) | 24 | 19(16) | Dyspnea, Sternal pain | Survive |

Abbreviations: HTN, hypertension; DM, diabetes; COPD, chronic obstructive pulmonary disease; CHB, chronic hepatitis B; GIST, gastrointestinal stromal tumor; HFNC, high-flow nasal cannula oxygen therapy; NIV, non-invasive ventilation; NP, nasal prongs; PP, prone position; LP, lateral position; CT, computed tomography.
\*Interval days of PP and/or LP sessions after the baseline.

The mean respiratory rate (RR) for all patients decreased from 28.4±3.5 breaths/min to 21.3±1.3 breaths/min after various durations of PP and/or LP sessions. Evaluation of CT results using the score system showed improvement in all patients, with a mean score decreasing from 41.8±19.2 to 29.3±16.8. For all patients, no new lesion was identified in the CT results (Table 2, Figure 2). No further deterioration occurred and no invasive intubation was required for all patients. All 25 patients survived. By March 17, 2020, all 25 patients have changed from severe to mild. By April 16, 2020, all patients have been discharged. Further follow-up up to May 3, 2020 indicated that no patients relapsed.

**Figure 2:**
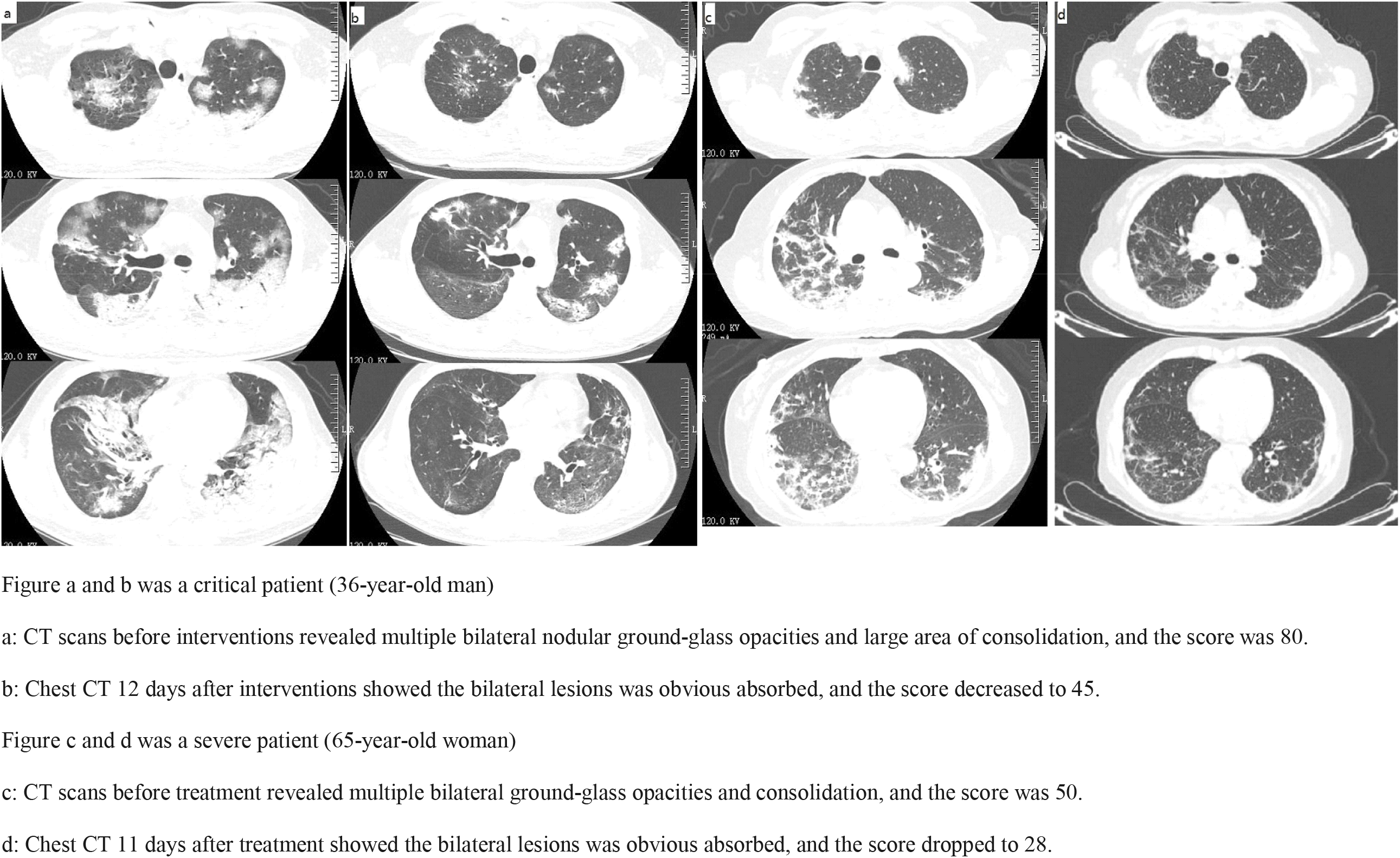
CT images of lungs from two patients and scores evaluated according to the scoring system

## Discussion

One of the pathophysiological features of ARDS in severe patients is lung inhomogeneity.^14^ Because of this, any delay in treatment will further cause reduced lung compliance, ventilation-perfusion imbalance, and decreased lung volume, which can lead to critical situations such as severe hypoxemia. According to the results of the autopsy of critical COVID-19 patients in Wuhan, China, main pathological features include exudation, infiltration of macrophages, and fibrosis in the lungs. The presence of mucous plugs with fibrinous exudate in respiratory tracts including distal regions is a distinctive feature of COVID-19.^15^ The clinical data of patients, however, showed that COVID-19 will not cause “typical” ARDS symptoms, which might partially explain the poor response of critical COVID-19 patients to lung recruitment and positive end-expiratory pressure (PEEP) associated with mechanical ventilation.^16^ Therefore, oxygen therapy and mechanical ventilation cannot fully resolve the pathology of ARDS caused by COVID-19 due to their working mechanisms. The use of prone position, however, can mobilize copious secretions and promote forward drainage toward the central airways for clearance.^17^ PP also improves lung inhomogeneity, redistribution of perfusion^18^, lung compliance, recruitment-to-inflation ratio, and lung recruitment.^19,20^ Therefore, for severe ARDS caused by COVID-19, many clinical guidelines^8-11^ recommend prone position for critical patients on mechanical ventilation.

Most studies on prone position were focusing on mechanically ventilated patients with severe ARDS. Few studies ^21,22^ explored the effect of lateral position as well. In recent years, some studies ^23,24^ reported the efficacy of prone position in combination with HFNC or NIV in severe and moderate ARDS and suggested that these interventions can help prevent intubation in some patients. During the treatment of patients with COVID-19, we’ve been thinking whether we can apply PP and/or LP as an early intervention in managing severe and even critical patients with severe hypoxemia and/or hypercapnia. We also noticed that most clinical guidelines for ARDS do not clearly recommend or suggest applying prone position or lateral position in patients with mild ARDS. One possible reason for this lack of clinical guidelines suggesting the use of PP and/or LP could be because most of the patients with severe ARDS, admitted into the ICU, require intubation. Also, patients with mild to moderate symptoms rarely get admitted into ICU. In previous studies on prone ventilation in treating ARDS patients, patients included in the studies have various diseases, clinical courses, or severity, thereby making it hard to draw conclusions that are comparable due to various differences.^25^ This time, despite of two time-related phenotypes of COVID-19 pneumonia,^26^ as our patients are all COVID-19 patients, their pathological abnormalities are almost the same, it enables us to better observe and analyze the ARDS associated with COVID-19.

Based on the clinical practices implemented during the study period, we found that after using PP and LP, patients’ oxygenation was improved. With the rapid increase of severe and critical patients who require oxygen therapy or higher respiratory support, current global medical resources are insufficient and might not provide adequate support. As a result, if we wait until patients deteriorate to a critical stage, where they require mechanical ventilation and prone position, significant medical resources will be needed and utilized (e.g. ventilators, staff for intubation, proper positioning and monitoring, regular suctioning, etc). This will increase the workload of the medical staff. In addition, due to the strong transmission of COVID-19, proper personal protective equipment (PPE) is in huge demand. The inability to match the supply with the demand associated with the global shortage of PPE will, inevitably, increase the risk of exposure for clinical staff. If we consider use PP and/or LP in combination with oxygen therapy or NIV earlier in their stage of disease development, the patients might tolerate better and perform PP and/or LP without sedation and nursing support. Although in this study, the PaO2/FiO2 value only indicated the improvement of patients’ status at one particular moment post PP and/or LP sessions, the CT images also showed the improvement from another perspective. No new lesions were identified and no severe adverse events occurred. It should also be noted that all the clinical staff of our team have cover-all suits as proper PPE during procedures and daily practice.

In terms of daily PP sessions for treatment of patients with severe ARDS, prior studies^27,28^ have provided various recommendations around 6-16 hours with the longest session lasting upto 20 hours. Another meta-analysis^29^about using prone position on patients with moderate to severe ARDS found that prone ventilation for at least 12 hours per day can improve patients' oxygenation and prognosis. The patients in the meta-analysis, however, were all on mechanical ventilation and sedated. The patients in our study, on the contrary, were conscious and used active range of motion. In our actual clinical practice, we further tailored daily PP and/or LP sessions for all patients based on their tolerance and response. For patients with severe ARDS, the requirement was to use at least 4 hours of prone position plus any other active position. For patients who were recovering quickly, the requirement was to use at least 10 hours of prone position. For critical patients, depending on their situation, the hours of each session varied. The results showed that critical patients tolerated a mean daily PP session of 6 hours during study period. One particular patient even tolerated one PP session of 16 hours.

As this observational study was done during the outbreak, it has some limitations. First, there were limited medical resources which impacted the oxygen support provided to patients. It should also be noted that the study was conducted in China, and thereby the environment and resources might be different from other countries. Second, the sample size is small considering the nature of an observational study. Although we noticed that a single case report^30^ and a pilot study^31^ have reported on similar strategy used in this study, our study reported on the clinical data of severe and critical patients during a long period of treatment and follow-up. Thirdly, some specific data about tests were missing. However, the limited findings still indicated positive effects of using PP and LP in managing patients with COVID-19. It would require large scale prospective clinical trials to explore the generalizability of these interventions among other COVID-19 patients in other countries and/or environments. In conclusion, using prone position and lateral position among patients at the early stage of the disease required less medical resources, and the outcome of this study suggested that the risks of generating new lesions and/or complications were low. Therefore, it might be beneficial to consider this as an early intervention, especially under the context of pandemic and limited medical resources.

## Data Availability

No data avilabale. All data was retrieved from the medical records, including demographic features,
clinical features, laboratory findings and chest CT images.

## Acknowledgments

We thank all patients involved in the study.

## Disclosure

All authors have declared no conflicts of interest.

## Author contributions

L.B., SL.X., YP.G., conceived the study. W.D., J.F., collected the data. HM.Q., P.Z., evaluated the CT images. W.D., SL.X, J.F., Y.D., JC.Z., conducted data analysis. SL.X., W.D., L.B., drafted the manuscript. YP.G., L.B., SL.X. and W.D revised the draft of the manuscript. All authors read and approved the final manuscript.

## Funding

The authors received no financial support for the research, authorship, and publication of this article.

## Notes

### Competing Interest Statement

The authors have declared no competing interest.

